# Details of “conversion therapy” practices and concordance with legislative definition: Findings from a non-probability community-based survey in Canada, 2020

**DOI:** 10.1101/2021.11.15.21266353

**Authors:** Travis Salway, Amrit Tiwana, Julia Schillaci-Ventura, Sarah Watt, Erika Muse, Florence Ashley

## Abstract

**Background:** “Conversion therapy” practices (CTP) refer to a heterogeneous set of treatments and activities that share the common goal of suppressing, “repairing,” or otherwise preventing expressions of transgender, lesbian, gay, bisexual, or queer identities. This study aimed to elicit details from those with direct experience and assess concordance between CTP experiences and the definition of CTP included in federal legislation aiming to deter CTP.

**Methods:** We conducted an anonymous online survey of adults (>18 years) in Canada between August 18 and December 2, 2020. Participants were recruited through social media, CTP survivor networks, and word-of-mouth.

**Results:** Of 51 respondents with direct experience of CTP, 16 were transgender, 34 were cisgender. Most respondents lived in Alberta, British Columbia, Ontario, or Quebec. 49% experienced CTP in a licensed healthcare provider office, 45% at a faith-based organization, and 36% at an unlicensed counselor office (categories not mutually exclusive). Age at first CTP experience ranged from 2 to 44 years of age (mean: 17.8 years; median: 17 years). Respondents experienced CTP for <1 year to 33 years (mean: 4.6 years; median: 2 years). Fifty percent of those with direct experience of CTP indicated that the proposed legislative definition of CTP did not fully encompass their personal experience.

**Interpretation:** Results from this Canadian survey of adults with experience of CTP indicate that legislative attempts to ban CTP must be expanded (e.g., to include adults and practices that do not concord with previously drafted definitions) and paired with other prevention efforts.

## Introduction

“Conversion therapy” practices (CTP) refer to a heterogeneous set of treatments and activities that share the common goal of suppressing, “repairing,” or otherwise preventing expressions of transgender, lesbian, gay, bisexual, or queer identities.^1,2^ CTP continue to occur throughout the world, despite denouncements by numerous health professional bodies, including every major medical and psychological association in North America.^3^ In the US and Canada, an estimated 4-18% of sexual and gender minorities have experienced CTP.^4–9^ The psychological consequences of experiencing CTP are pronounced and well-demonstrated. For instance, research suggests that approximately 30% of those who experience CTP attempt suicide at least once.^10–13^

In this context, numerous jurisdictions have begun enacting legislative bans on CTP in recent years. To date, bans have been passed in five countries (i.e., Malta, Brazil, Ecuador, Taiwan, and Germany)^3^, twenty US states^14^, and five Canadian provinces and territories.^15^ These bans differ with regard to their definition of CTP, inclusion of CTP targeting gender identity and/or expression, settings/practitioners targeted, type of sanction, and age groups protected.^1,3^ For example, bans passed in many US states are focused on licensed health professionals treating minors^16^, while a recent ban passed in the Australian state of Victoria extends their ban to religious practitioners and protects adults as well as youth^17^.

In March 2020, the Canadian federal government introduced Bill C-8 (later re-introduced as Bill C-6, October 2020), *An Act to Amend the Criminal Code (conversion therapy)*.^18^ Bill C-6 (now defunct) would have prohibited non-consensual CTP for people of all ages, any form of CTP for minors, removing a child from Canada to undergo CTP abroad, receiving financial or material benefit from CTP, and advertising CTP.^19^ At its introduction, Bill C-6 defined CTP as “a practice, treatment or service designed to change a person’s sexual orientation to heterosexual or gender identity to cisgender, or to repress or reduce non-heterosexual attraction or sexual behaviour.” To be most effective, attempts to prevent CTP—including legislative bans—need to be informed by a current, empirical understanding of the forms, settings, ages, and monetary exchanges associated with CTP. To date, few studies have explored these important details, and only one has been conducted in Canada.^8,20–22^ International studies show that CTP occur across a wide range of settings and practitioners, suggesting that effective CTP bans must account for the varied ways in which CTP are framed and advertised.^3,20,21^

In order to directly inform the Canadian Parliament’s amendments to Bill C-6, we conducted a rapid survey of people in Canada with direct and indirect experience of CTP. Our primary objectives were to elicit details from those with direct experience and assess concordance between CTP experiences and the definition of CTP in Bill C-6. We additionally queried those with indirect experience of CTP (i.e., nearly experienced CTP or know someone who experienced CTP) about their knowledge of ongoing CTP in Canada.

## Methods

### Sample

We conducted an anonymous, open, online survey between August 18 and December 2, 2020. To be eligible, participants had to have: experienced CTP, nearly experienced CTP, or known someone who has experienced CTP; been 19 years of age or older; and resided in Canada. The study was promoted through e-mails and social media advertisements targeting existing networks (e.g., previously identified people with direct experience of CTP^23^) and partner community-based organizations from across Canada. Prospective participants were directed to a university webpage describing the purpose and nature of the study. Respondents identified that they heard of the study through word-of-mouth (37%), Twitter or Facebook (35%), Two-Spirit, lesbian, gay, bisexual, transgender, and/or queer community organizations (15%), and participation in an in-depth interview with our research team^23^ during the spring/summer of 2020 (13%). A total of 108 individuals began the survey, of which 70 (65%) were eligible and completed questions related to our primary objectives, thereby forming the analytic sample for the present report. Participation was voluntary, and all participants provided informed consent prior to completing the questionnaire. No incentives were provided.

### Measures

Questionnaire content was developed in consultation with CTP survivors and people working to prevent CTP or reduce harms associated with CTP in Canada. Most questions were created *de novo*, given the lack of existing measures; however, some questions were adapted from surveys of people with CTP experience conducted in other countries.^11,20^ To determine eligibility, respondents were asked: “How would you describe your experience with conversion therapy (i.e., structured activity to deny or suppress your LGBTQ2 identity)? Please check all that apply.” Respondents were classified as having direct experience if they selected “I have experienced conversion therapy” or “I think that I experienced conversion therapy, but I’m not sure.” Respondents were classified as having indirect experience if they selected “I nearly experienced conversion therapy, I considered going,” “I nearly experienced conversion therapy, someone suggested that I go (e.g., a parent/guardian, religious leader, teacher, counselor, healthcare provider),” or “I know someone who experienced conversion therapy.”

Respondents provided socio-demographic characteristics, including sex assigned at birth, gender identity, sexual orientation, age, racial/ethnic identity, and province of residence. We classified respondents as transgender if they reported a gender identity that differed from their sex assigned at birth and cisgender if they reported a gender identity that corresponded with their sex assigned at birth. Adaptive questioning was used. Respondents who indicated that they had direct experience were asked about: nature of CTP including targeted characteristic (sexual orientation, gender identity, or gender expression), religious identity at time of CTP, CTP setting, presence of others during CTP, gender(s) of others present also undergoing CTP, payment for CTP, and age and duration of CTP; preferred supports for healing from CTP; and, concordance between definition of CTP in Bill C-6 and personal experience, specifically, “Does the proposed definition of conversion therapy in Bill [C-6] encompass your experience with conversion therapy?: very much; somewhat; undecided; not really; not at all”. Respondents with indirect experience were asked about number of contacts who experienced CTP and known advertising mechanisms for CTP. The online questionnaire spanned 12 pages.

### Data analysis

Analyses were descriptive, using counts/proportions and measures of range and central tendency (mean, median), as appropriate. Density plots were used to examine the distribution of age at first CTP experience and duration of CTP experiences. Statistical analyses were completed using RStudio version 1.4.1106 (R version 4.0.4). Where personal CTP experiences were discordant with the definition in Bill C-6, we elicited open-ended explanations. These responses were then organized by the first author using conventional content analysis, in which codes are derived inductively.^24^

### Ethics approval

The protocol for this study was approved by the Simon Fraser University Research Ethics Board (study 2019s0394).

## Results

### Summary of sample

A total of 51/70 (72.9%) respondents reported direct experience with CTP, 41/70 (58.6%) reported indirect experience, and 22/70 (31.4%) reported both direct and indirect experience. Of those with indirect experience, 36/41 (87.8%) knew someone who had experienced CTP, 7/41 (17.1%) nearly went to CTP because someone suggested it, and 2/41 (4.9%) nearly went without any external pressure to attend (categories not mutually exclusive). Respondents were 20-90 years of age (mean: 37.6 years; median: 33 years). Among those with any direct experience (n=51), 31% were transgender; among those with *only* indirect experience (n=19), 48% were transgender (**Table 1**).

**Table 1.**
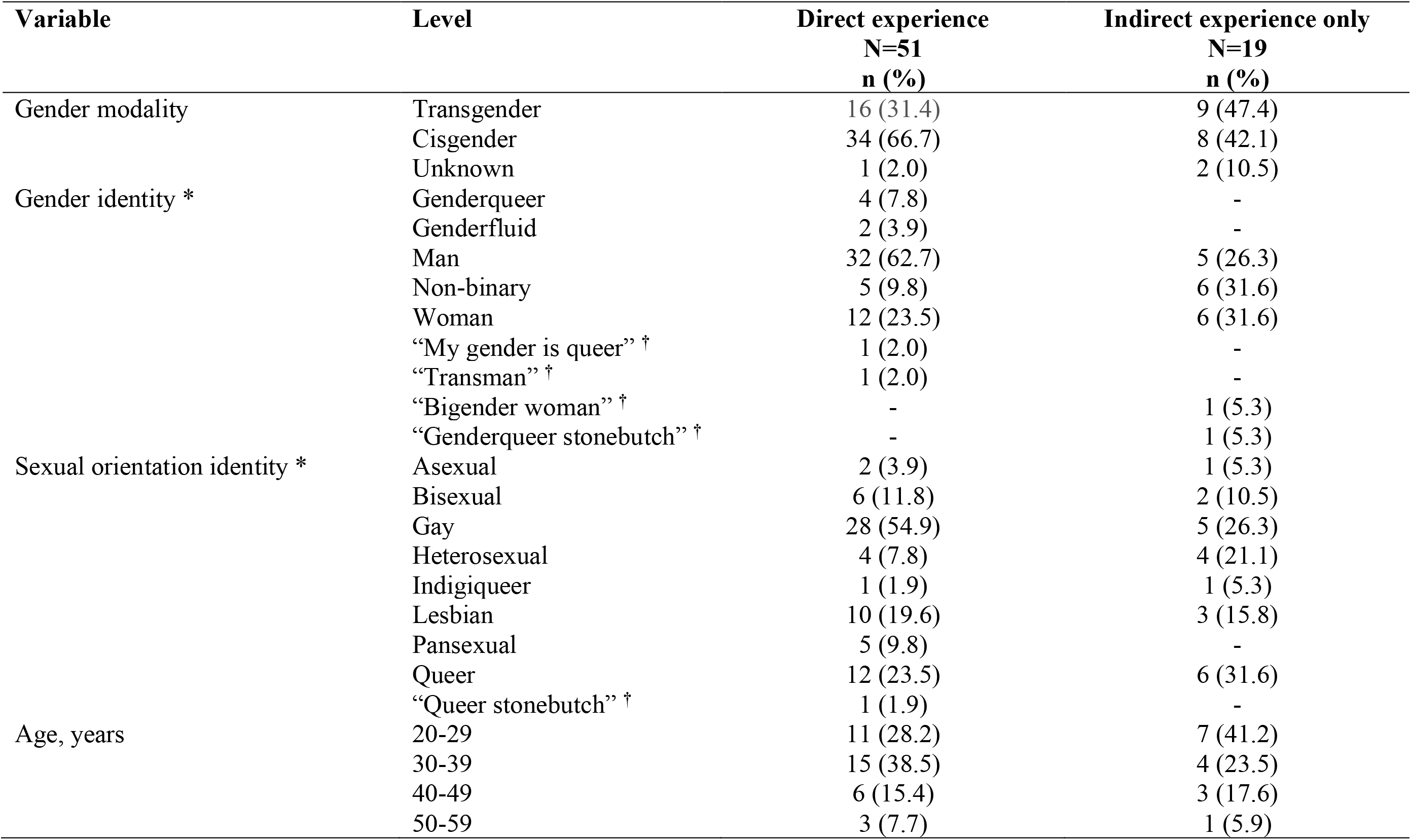
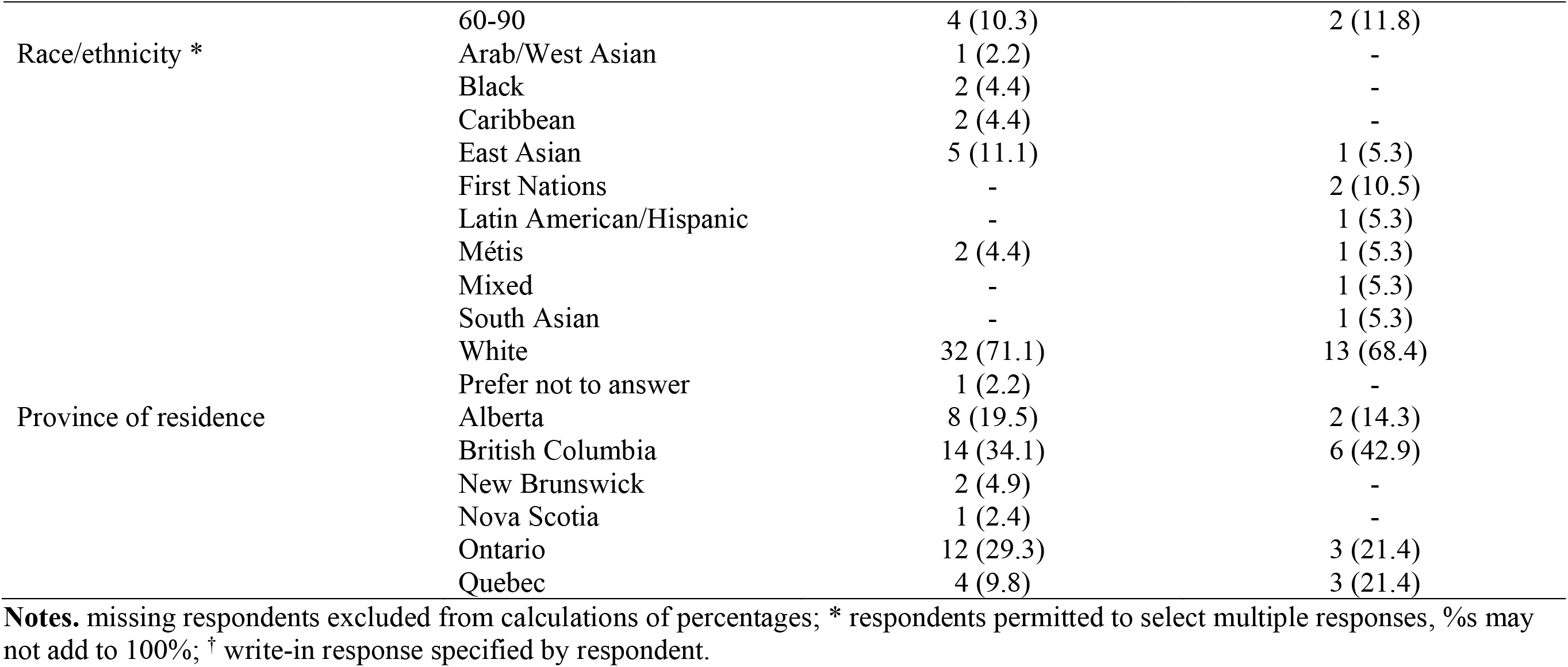
Characteristics of respondents to a Canadian survey regarding direct and indirect experiences with “conversion therapy” practices, September – December 2020

### Details of direct experiences

Among transgender respondents with direct experience, 21.4% experienced CTP targeting only gender identity, while most experienced CTP targeting some combination of gender identity, gender expression, and sexual orientation (**Table 2**). Among cisgender respondents, most (77.4%) experienced CTP targeting only sexual orientation. No single setting was reported by more than half of the respondents with direct experience of CTP, and 49% experienced CTP by a licensed healthcare provider. Age at first CTP experience ranged from 2 to 44 years of age (mean: 17.8 years; median: 17 years) (**Figure 1**). Respondents experienced CTP for <1 year to 33 years (mean: 4.6 years; median: 2 years) (**Figure 2**).

**Table 2.**
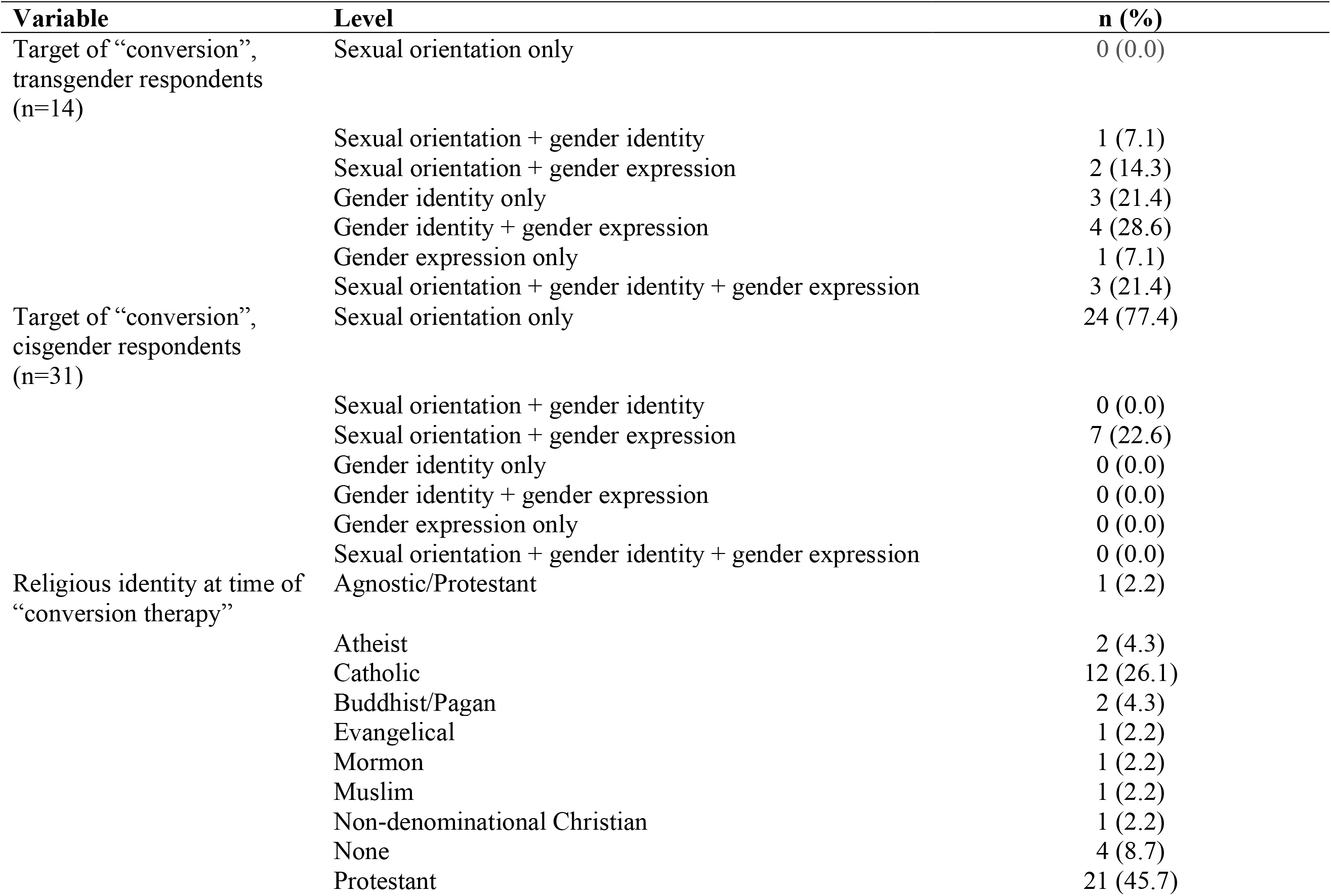
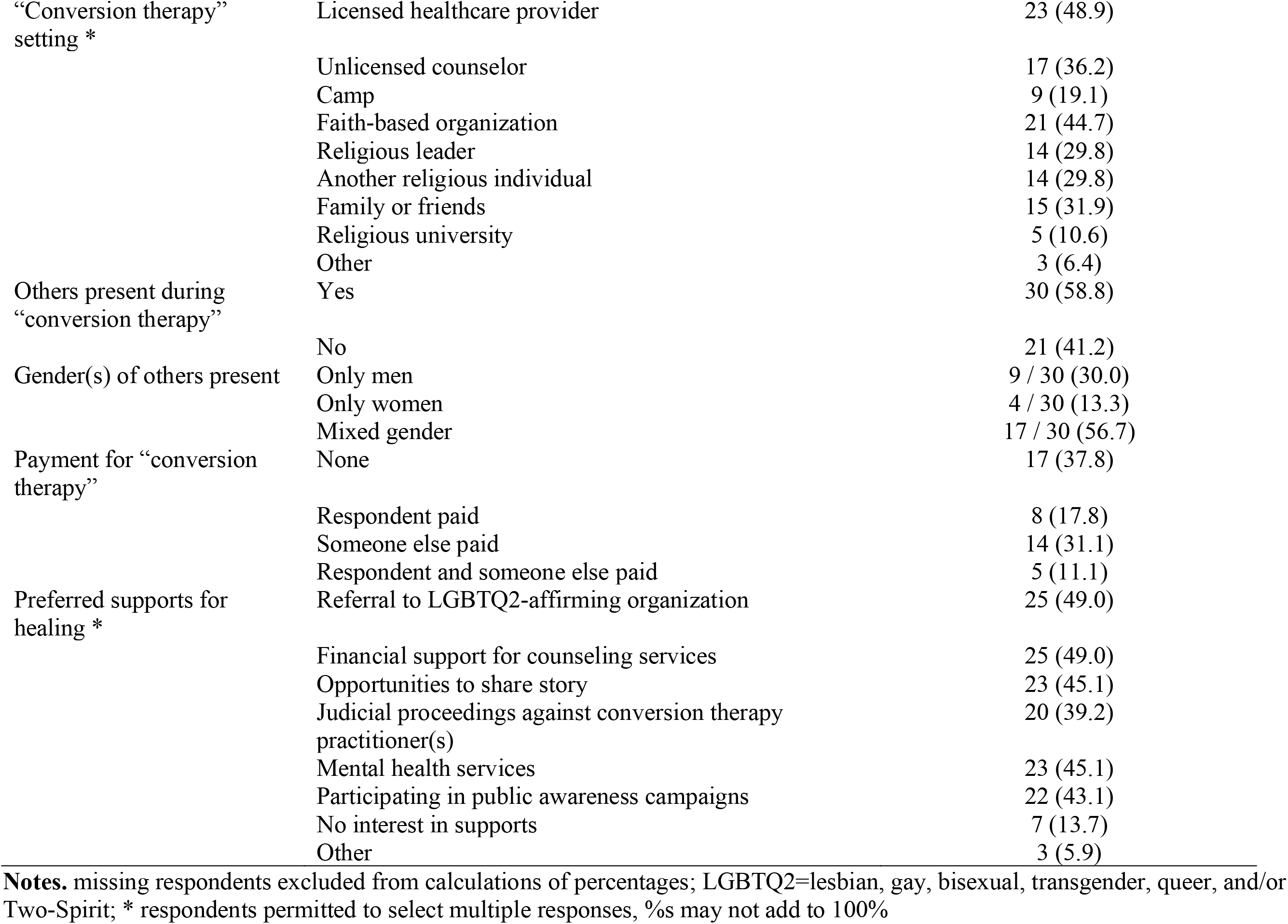
Details of direct experiences with “conversion therapy” practices, reported in a Canadian survey, September – December 2020, N=51

**Figure 1.**
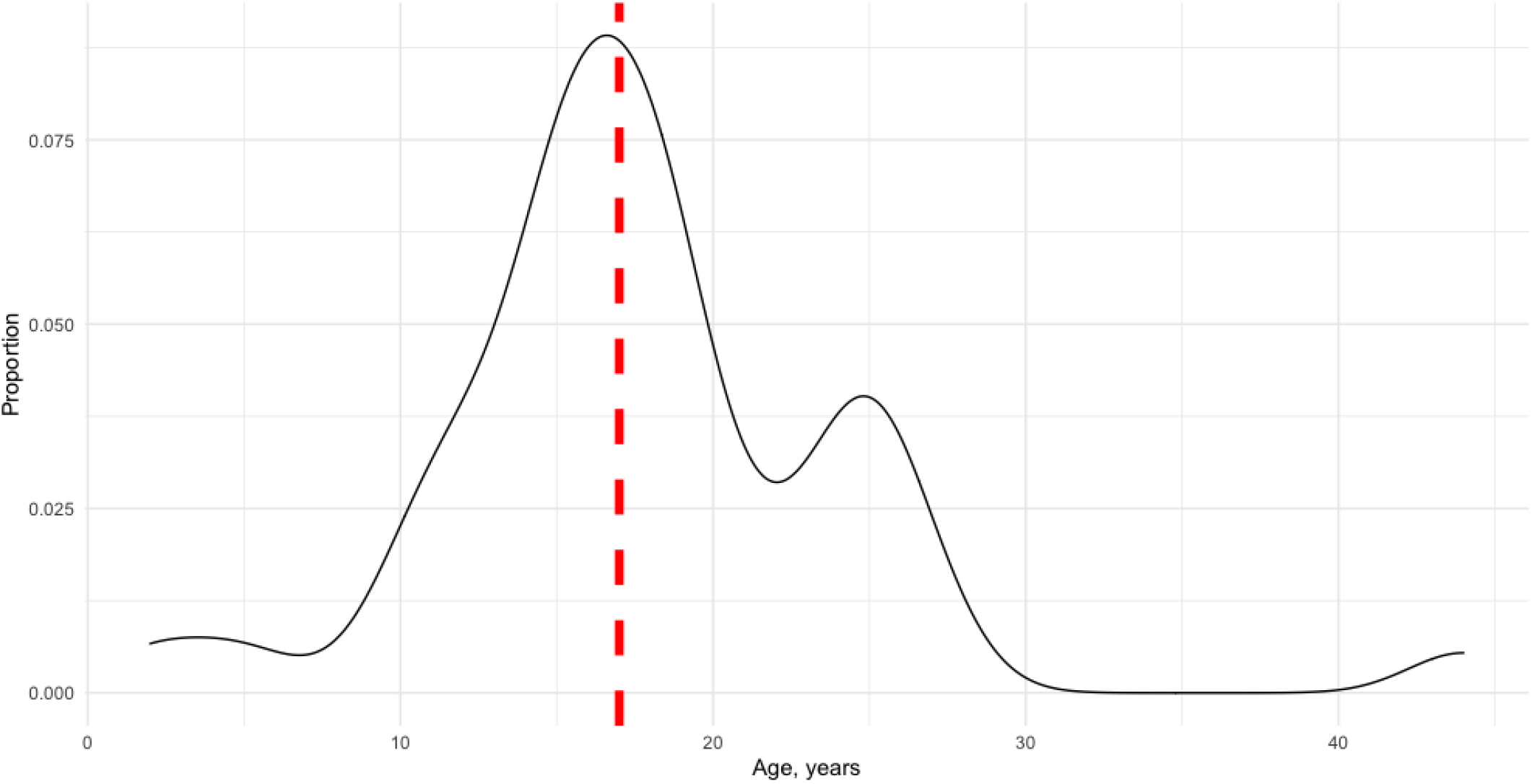
Density plot of age at first “conversion therapy” experience among respondents to a Canadian survey, September – December 2020, N=43 (vertical red line = median)

**Figure 2.**
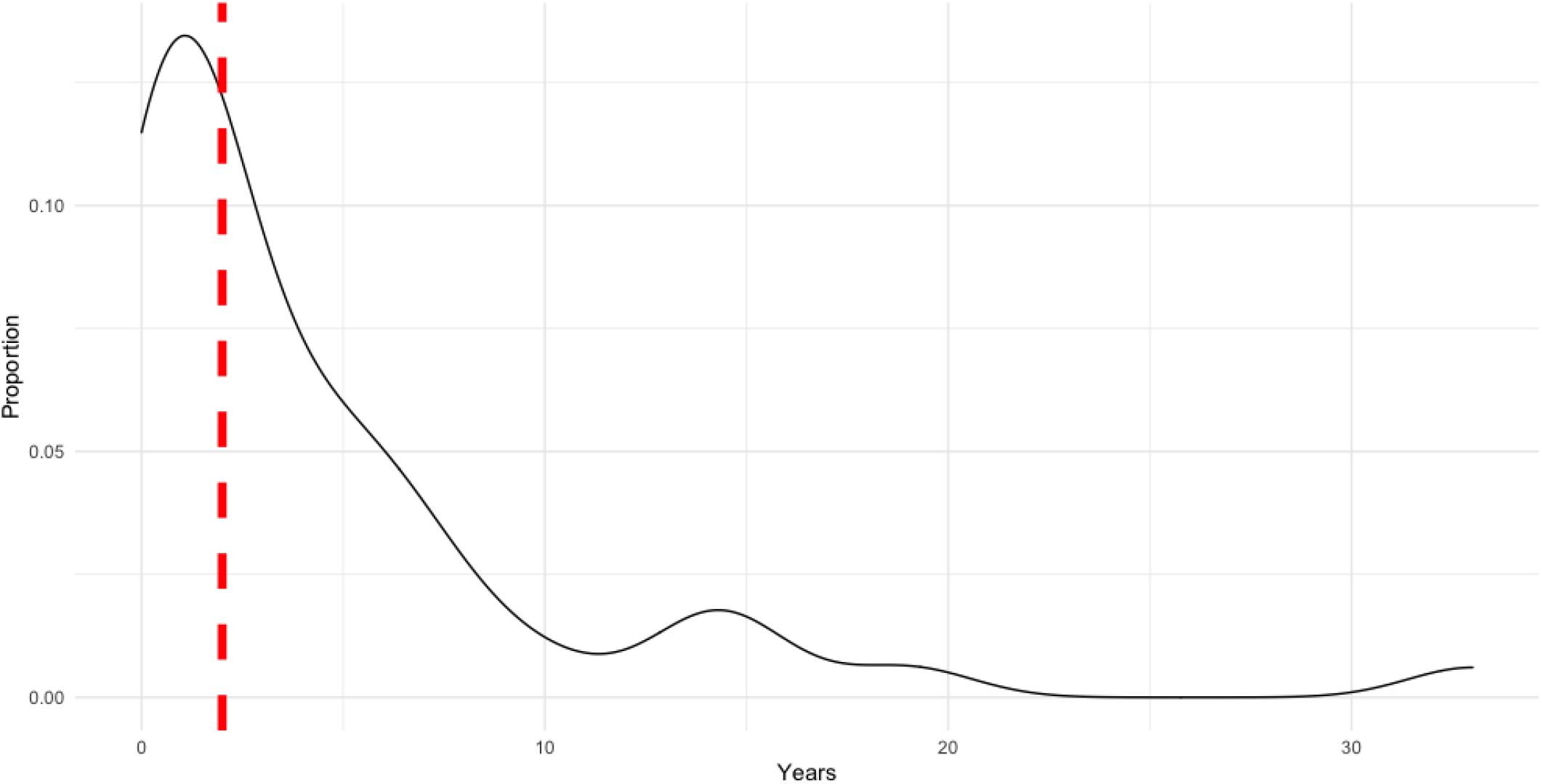
Density plot of duration of “conversion therapy” experience among respondents to a Canadian survey, September – December 2020, N=43 (vertical red line = median)

### Details of indirect experiences

A total of 43 respondents reported personally knowing at least one person who had experienced CTP (excluding themselves), and the number of contacts with CTP experience ranged from 1 to 100+ (mean: 18.4, median: 5). Collectively, these 43 individuals knew a total of more than 793 people who have experienced CTP. Respondents were aware of the following methods used by conversion therapy practitioners to advertise their services (n=41 complete responses): word of mouth (61.0%); sermons or talks at faith-based institutions (43.9%); community bulletin boards (19.5%); internet or social media advertisements (31.7%); conference presentations (24.4%); published books or brochures (22.0%); and referrals from healthcare providers (22.0%).

### Concordance between legislative definition and direct experiences

Fifty percent of those with direct experience of CTP indicated that the proposed definition of CTP in Bill C-6 encompassed their personal experience, while 29.2% indicated that it somewhat encompassed their experience, and 20.8% indicated that it did not encompass their experience. Explanations provided by those whose direct experience of CTP was discordant with the definition in Bill C-6 fell into three categories (**Table 3**): (1) exclusion of trans conversion practices; (2) experience of a practice that was not defined by “changing” or “repressing” a sexual orientation or gender identity; and (3) experience that was not a circumscribed practice or treatment.

**Table 3.**
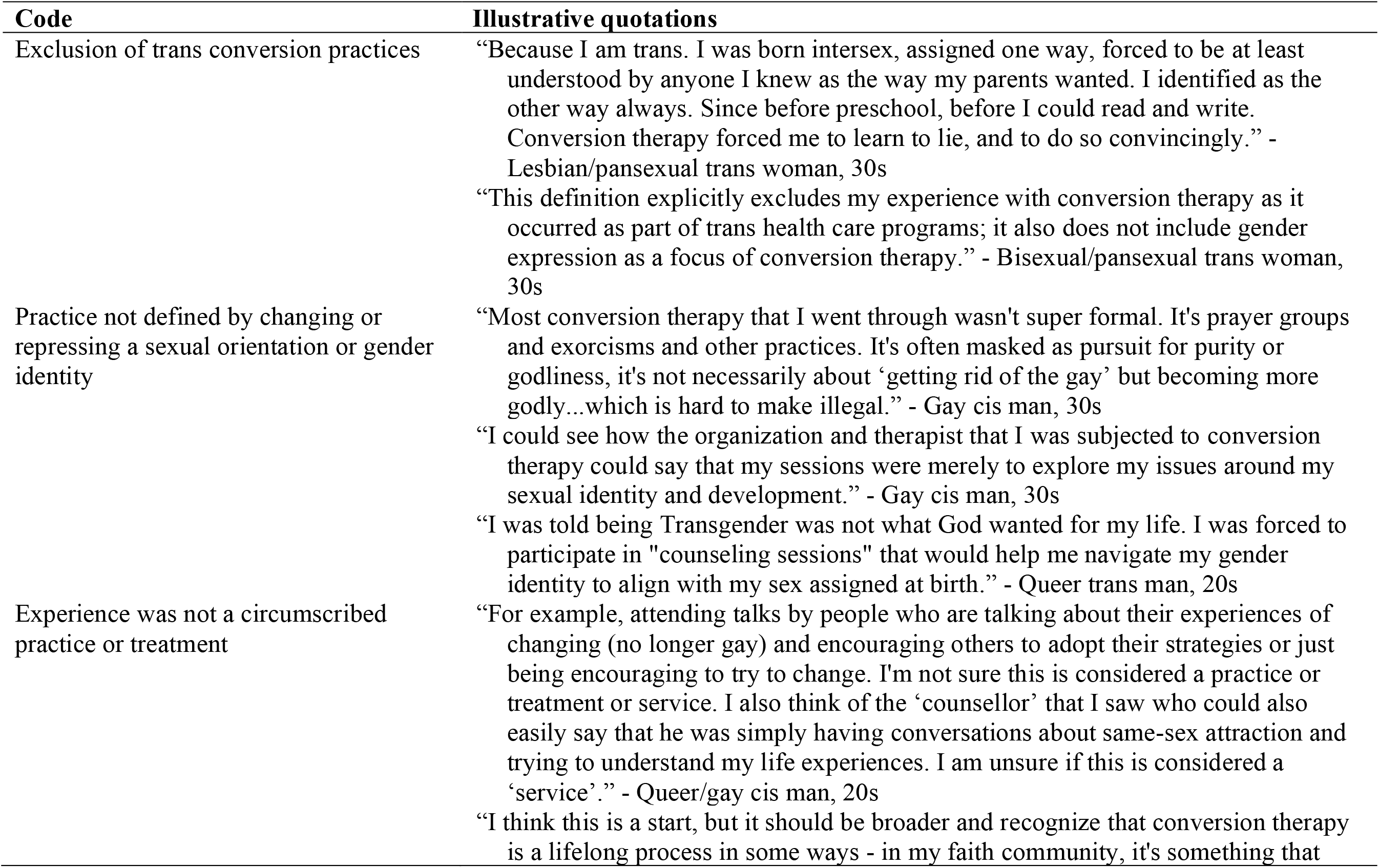
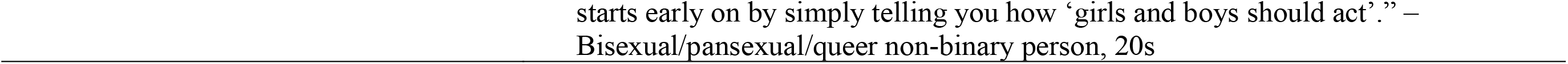
Explanations provided by those with direct experience of “conversion therapy” practices in Canada, regarding discordance between personal experience and legislative definition of “conversion therapy”

## Discussion

This rapid, non-probabilistic survey of adults in Canada with direct or indirect experiences of so-called “conversion therapy” practices yielded detailed insights about the nature of CTP, which remains a topic of debate and concern among legislators in Canada, and internationally. Most importantly, our findings point to important limitations in the proposed Canadian ban on CTP (Bill C-6, 2020-21) and suggest that more than half of those surveyed with direct experience would not have been protected by this bill, on the basis of its definition of CTP and/or the age at which they began CTP.

Bill C-6 limited its protection to minors in Canada. We found, however, that half of the survey respondents began CTP as adults (**Figure 1**). This aligns with findings from a US survey (2008-09), in which the median age at first experience of CTP was 20 years,^20^ while suggesting a slightly older age distribution of age at CTP initiation than that identified in a recent Canadian survey (2019-20), in which 28% first experienced CTP at 20+ years of age.^8^ With regard to settings, as with previous studies^7,8,20,21^, our results demonstrate that CTP in Canada is not relegated solely to the domain of religious practitioners. Nearly half of the survey respondents experienced CTP with a licensed healthcare provider, and others still experienced CTP with an unlicensed counselor or with family or friends. We estimated that 38% experienced CTP in which no payment was made to the practitioner—an important detail given that Bill C-6 would have prohibited “receiving a financial or other material benefit from the provision of conversion therapy.”^19^

By conducting this study concurrent to the Canadian Parliament’s introduction and debate of Bill C-6, we had a unique opportunity to present survey respondents with the definition of CTP used in Bill C-6, in order to gauge the degree to which their direct experiences corresponded to the federal legislators’ proposed definition.^19^ Half of these respondents indicated some degree of discordance between the definition in Bill C-6 and their personal experiences.

After the introduction of Bill C-6, CTP survivors, researchers, and activists published an open letter informing the federal government that the wording of Bill C-6 was insufficient to capture CTP that target trans people.^25^ Consistent with this critique, we found that trans survey respondents had experienced CTP as part of government sanctioned healthcare programs which were not designed to “change a person’s… gender identity cisgender.” Others pointed to the absence of language regarding gender expression as a common target for CTP; indeed, 57% of transgender respondents and 23% of cisgender experienced CTP that included gender expression as a targeted characteristic (**Table 2**).

Other respondents outlined ways in which the CTP they experienced was not advertised or defined as “conversion therapy” but rather as “prayer groups,” “pursuits for purity or godliness,” “sessions… to explore issues around sexual identity and development,” or simply “counseling sessions” (**Table 3**). These critical reflections highlight an inherent limitation to attempts to prevent CTP by defining it as “a practice, treatment or service designed to *change* a person’s sexual orientation… or gender identity.” Thus, we have previously argued that the defining feature of CTP is not an attempt to “change” or “convert” but rather “the goal of avoiding acceptance and acknowledgement of lesbian, gay, bisexual, transgender, queer, and Two-Spirit lives as compatible with being healthy and happy”.^2^ Finally, some survey respondents described an experienced that was not a circumscribed practice or treatment but rather a series of conversations—often with multiple people—that consistently doubted or dismissed the viability of being LGBTQ2. We have previously described these less-well delineated experiences as sexual orientation and gender identity and expression change efforts; curtailing these broader efforts will require interventions and policies that operate alongside legislative bans, which target more circumscribed CTP.^26^

### Limitations

Our survey was limited by its relatively small sample size, insufficient data to permit detailed analyses by intersecting social characteristics (notably gender, race/ethnicity, and geography), and lack of contextual information to interpret some findings. For example, we are unable to discern the nature of the CTP reported by one participant as beginning at age 2 years. While we sought to provide participants with precise definitions, it is possible that some participants may have reported experiencing CTP when in fact their experience was more akin to unorganized pressure to change or suppress an LGBTQ2 identity, or other less formalized expressions of cissexism or heterosexism. Finally, we focused this study on adults, 19+ years of age, and thus are unable to represent the experiences of youth experiencing contemporary CTP. In light of these limitations, the results of this survey should be confirmed and refined using a larger sample. Acknowledging that some who experience CTP are as a result disconnected from LGBTQ2 communities^23^, future studies should aim to sample using broader recruitment mechanisms.

### Conclusion

Experiences of CTP in Canada are heterogeneous, making it difficult to arrive at a unifying definition. Accordingly, half of the respondents to this 2020 survey who had direct experience of CTP indicated that a recent legislative definition of CTP failed to encompass their experiences. Survey respondents with direct experience of CTP also reported a prevalent desire for supports to aid in the healing and recovery from CTP, including provision of financial supports, access to affirming counseling, and judicial proceedings against conversion therapy practitioners. In concert, these findings point to the need for a multi-faceted approach to CTP prevention.^27^ We have previously estimated that >50,000 people in Canada have experienced CTP^8^; resources should be mobilized by governments posthaste to provide these supports as reparations for the harm inflicted upon them. The definition of CTP in legislative bans should be carefully developed through consultations with those with lived experience. Finally, legislative attempts to ban CTP should be paired with other CTP prevention efforts, including those that reduce the demand for CTP, i.e., through education, policy, and other social interventions that affirm LGBTQ2 identities.

## Data Availability

Given the highly sensitive nature of the data collected, the dataset for this study is not publicly available.

## Acknowledgments

This study was supported by Andrew Beckerman and the Victoria Foundation. We are grateful to the following individuals who provided feedback on draft versions of the questionnaire and/or who helped to promote the survey: A.J. Lowik, Beth Carlson-Malena, David Kinitz, Elisabeth Dromer, Geron Malbas, Keith Murray, Kiffer Card, Matt Ashcroft, Michael Kwag, Nicholas Schiavo, and Trevor Goodyear. We additionally wish to thank Stephen Juwono for his assistance with data cleaning.

